# Optimal health and economic impact of non-pharmaceutical intervention measures prior and post vaccination in England: a mathematical modelling study

**DOI:** 10.1101/2021.04.22.21255949

**Authors:** Michael J. Tildesley, Anna Vassall, Steven Riley, Mark Jit, Frank Sandmann, Edward M. Hill, Robin N. Thompson, Benjamin D. Atkins, John Edmunds, Louise Dyson, Matt J. Keeling

## Abstract

**Background:** Even with good progress on vaccination, SARS-CoV-2 infections in the UK may continue to impose a high burden of disease and therefore pose substantial challenges for health policy decision makers. Stringent government-mandated physical distancing measures (lockdown) have been demonstrated to be epidemiologically effective, but can have both positive and negative economic consequences. The duration and frequency of any intervention policy could, in theory, could be optimised to maximise economic benefits while achieving substantial reductions in disease.

**Methods:** Here we use a pre-existing SARS-CoV-2 transmission model to assess the health and economic implications of different strengths of control through time in order to identify optimal approaches to non-pharmaceutical intervention stringency in the UK, considering the role of vaccination in reducing the need for future physical distancing measures. The model is calibrated to the COVID-19 epidemic in England and we carry out retrospective analysis of the optimal timing of precautionary breaks in 2020 and the optimal relaxation policy from the January 2021 lockdown, considering the willingness to pay for health improvement.

**Results:** We find that the precise timing and intensity of interventions is highly dependent upon the objective of control. As intervention measures are relaxed, we predict a resurgence in cases, but the optimal intervention policy can be established dependent upon the willingness to pay (WTP) per QALY loss avoided. Our results show that establishing an optimal level of control can result in a reduction in net monetary loss of billions of pounds, dependent upon the precise WTP value.

**Conclusions:** It is vital, as the UK emerges from lockdown, but continues to face an on-going pandemic, to accurately establish the overall health and economic costs when making policy decisions. We demonstrate how some of these can be quantified, employing mechanistic infectious disease transmission models to establish optimal levels of control for the ongoing COVID-19 pandemic.

## INTRODUCTION

In late 2019, the first cases of an unknown respiratory infection began to emerge in the city of Wuhan, in Hubei province, China [1, 2]. In order to attempt to control the spread of the virus, countries around the world introduced a range of measures, including mandatory physical distancing, wearing of face coverings, restrictions on large gatherings and, in situations where case numbers were increasing in an uncontrolled manner, regional or nationwide policies that have included closure of schools, restrictions on travel and in extreme cases, stay at home orders [3].

In the UK, the first cases of COVID-19 were reported in the city of York on 31st January 2020. Cases began to rise in a concerning manner in March 2020 and on 12th March, the government policy moved from “containment” to “delay”, suggesting the imminent introduction of restrictions in order to flatten the peak and avoid the National Health Service (NHS) potentially being overwhelmed in the following weeks. On 20th March, a wave of restrictions were introduced, including the closing of all schools to children other than the vulnerable and those with key worker parents, and the closing of all pubs, restaurants and indoor leisure facilities. Finally, on 23rd March, the UK prime minister announced a national lockdown, in which all individuals had to stay at home and were only allowed out for essential shopping, healthcare, essential work if they could not work from home and one form of exercise per day.

As cases continued to decline during April, May and June 2020, mandatory controls were gradually relaxed in order to mitigate further economic harm. in England, schools opened to certain year groups from early June, whilst non-essential shops re-opened on 15th June and pubs and restaurants on 4th July, albeit with physical distancing measures in place in an attempt to control the spread of the virus. As society gradually opened up, cases began to rise again, slowly at first, but increasingly rapidly in September and October. Given the potential for a significant second wave of infection, there was a call from many stakeholders for a short “circuit breaker” lockdown in England, in order to stem the rise in cases and protect the NHS during the winter months. The UK government finally introduced a 4-week lockdown on 5th November in England, whilst Scotland, Wales and Northern Ireland introduced similar, short-term mandatory control policies during October and November in an attempt to keep the virus under control.

Much of the existing modelling literature on the pandemic has focused explicitly on the impacts of interventions that minimise the direct health impact of the COVID-19 pandemic, such as the number of individuals being admitted to hospital and/or dying from the disease [4–6]. However, it is important to note that there are non-health benefits and harms that can arise as a result of lockdown. From an economic perspective, long-term closures of the hospitality sector and non-essential shops could lead to wholesale closure of businesses, which will in turn lead to increased unemployment. There are additionally effects on tourism, transport and productivity. These disruptions can in turn lead to indirect health harms, such as potential delays in elective hospital procedures, mental health harm and reduced fiscal space for healthcare spending as a result of economic contractions. On the other hand, voluntary physical distancing in response to increases in COVID-19 cases and deaths plays a role that is at least as important as government-mandated controls in driving economic losses, particularly in high-income countries [7, 8]. As a result, judicious use of lockdown measures may ultimately hasten economic recovery [9, 10]. It is therefore important to consider the effect of any control policy on the overall economic cost of an outbreak, taking into account both positive and negative health and economic effects.

There is significant debate around enhanced physical distancing in both the UK and around the world with a focus on a perceived trade-off between averting COVID-19 cases and avoiding non-COVID loss of welfare [11, 12]. However, there has only been a limited exploration of this perceived trade-off in analysing the economic valuation of vaccines (REF in comment). To date no paper has explicitly explored the optimal level of physical distancing in the UK context using a mechanistic model of disease transmission, that can be used to estimate the impact of interventions on cases, hospitalisations and deaths, incorporating economic impact, though previous work has investigated potential health and economic impacts of vaccination [12]. In this paper, we analyse the effectiveness of different control scenarios (including more intensive long-term control or intermittent periods of fixed-term control) taking into account the positive impact on public health and the negative impact on the economy. We aim to inform optimal physical distancing policies that minimise a given mixed objective function of social welfare (that combines economic and public health losses) to inform policy in the UK, and provide an example of analytical approach to assessing the COVID policies more broadly.

We focus here on two scenarios. We first evaluate past UK policy, to illustrate the potential trade-offs between two physical distancing policies during the second wave in late 2020. We then extend this analysis to appraise different future approaches to physical distancing as vaccination is rolled out. In the first scenario, we explore the impact of introducing multiple “precautionary breaks” – short-term lockdowns of up to four weeks duration – would have had during 2020 and investigate the predictions of the cost of such a policy, dependent upon the intensity of control both within and outside these lockdown periods. We consider different lockdown period durations, intensity and timings, and calculate an overall cost for each measure. In the second scenario, we investigate the optimal relaxation policy to exit the 2021 lockdown, as dependent upon the pace of the vaccination campaign and the acceptable level of cost of the intervention policy.

To establish the COVID-related health impacts, we calculate the Quality Adjusted Life Year (QALY) loss for each scenario. Additionally, we calculate the estimated Gross Domestic Product (GDP) loss across this period and, by combining these measures, we determine an optimal scenario that is based upon monetising QALY losses using a societal willingness to pay (WTP) conversion factor. We apply the willingness-to-pay concept which can be used to evaluate direct costs incurred by health care services from a demand side perspective. This provides a way to measure the overall societal valuation of disease control, with an optimal policy determined based upon the policy maker’s WTP for health improvement. Our work provides a framework to facilitate decision making that takes into account the negative impacts that mandatory controls may have to economic productivity, as well as the value of positive health impacts that occur as a result of the reduction in the spread of disease.

## Methods

### The epidemiological model

In this paper we used a previously developed deterministic, age-structured compartmental model, stratified into five-year age bands [13]. It is important for age-structure to be incorporated into a model of this type, given that the risk of hospitalisation and death is highly dependent upon age. Transmission was governed through age-dependent mixing matrices based on UK social mixing patterns [14, 15]. The population was further stratified according to current disease status, following a susceptible-exposed-infectious-recovered (SEIR) paradigm, as well as differentiating by symptoms, quarantine and household status. We assumed therefore that susceptibles infected by SARS-CoV-2 would enter a latent state before becoming infectious. Given that only a proportion of individuals who are infected are tested and subsequently identified, the infectious class in our model was partitioned into symptomatic (and hence potentially detectable), and asymptomatic (and likely to remain undetected) infections. We assumed both susceptibility and disease detection were dependent upon age. We modelled the UK population aggregated to 10 regions (Wales, Scotland, Northern Ireland, East of England, London, Midlands, North East and Yorkshire, North West England, South East England, South West England), with each region modelled independently (i.e. we assumed no interactions occurred between regions).

A drawback of the standard SEIR ordinary differential equation (ODE) formulation in which all individuals mix randomly in the population is that it cannot readily account for the isolation of households. For example, if all transmission outside the household is set to zero in a standard ODE model, then an outbreak can still occur as within-household transmission allows infection between age-groups and does not account for local depletion of susceptibles within the household environment. We addressed this limitation by extending the standard SEIR models such that first infections within a household are treated differently from subsequent infections. To account for the depletion of susceptibles in the household, we made the approximation that all within household transmission was generated by the first infection within the household (see Supporting Information for further details).

## Model Solutions

### Scenario 1: Retrospective Analysis

For our retrospective analysis, we used the transmission model to simulate the spread of SARS-CoV-2 infection in the UK in the presence of none, one or two precautionary breaks. We ran a suite of simulations, varying the timing of the initial and the subsequent precautionary break, as well as the levels of control both within the precautionary break and during the intrinsic (inter-break) period; although we included a constraint that the level of control within a precautionary break must be at least as severe as the level of control outside the precautionary break. The earliest date we assumed the first precautionary break could occur was 1st July 2020. The minimum delay between the onset date of the first and second precautionary break was set to 4 weeks, based on the practical assumption that the UK government would be unlikely to implement two consecutive precautionary breaks within a short time period. We assumed each precautionary break was in place for 1, 2, 3 or 4 weeks. All simulations spanned February 2020 until the end of June 2021, although we calculated economic and health costs from June 2020 through to January 2021 (corresponding to the time period during which precautionary breaks could be enacted). Overall, we explored a wide set of potential future scenarios (approximately 50,000) where the placement of precautionary breaks and strength of controls were tunable parameters.

In our model, we considered the following variables when determining the optimal level and type of control:

i. The intensity of controls both within precautionary break periods and outside these periods. We measured this by a single parameter (*ϕ*, between zero and one) that combined the scale of any measures, public adherence and reactive public behaviour, in other words both mandated and voluntary distancing. When this parameter is zero, there are no restrictions or voluntary distancing, and thus additional impact on the economy, but transmission is maximal. When this parameter is one, there is very strong extent of physical distancing, comparable with the situation in the UK in early April 2020. Our constraint that the level of control within a precautionary break must be at least as severe as the level of control outside the break was enforced by ensuring the intensity of controls within break periods, *ϕ*_*PB*_, had a value equal to or greater than the intensity of controls outside of break periods, *ϕ*_*O*_ (i.e. *ϕ*_*PB*_ ≥ *ϕ*_*O*_). More details of the effect of *ϕ* upon transmission are given in the supplementary material.
ii. The duration of precautionary breaks. This was modelled as a fixed time interval (1, 2, 3 or 4 weeks), during which more severe intervention policies were introduced, compared to the intrinsic control measures. The duration of the break period(s) for a given simulation could be varied to establish the optimal duration of a fixed term precautionary break.
iii. The frequency and timing of precautionary breaks. We considered the impact of single and multiple breaks. We assumed that the intensity of each break was the same in a multi-phase strategy (though we acknowledge that, in practice, adherence could wane through time given repeated precautionary breaks).
iv. Limits on hospital occupancy. Given that the choice of policy may be influenced by the current number of individuals in hospital with COVID, we investigate the impact of a hospital occupancy threshold upon our choice of policy. We therefore consider hospital occupancy limits of 10,000 and 20,000 individuals and an alternate scenario where there is no limit on hospital occupancy.

### Scenario 2: Prospective Scenario

For the prospective scenario, we simulate the model forward from January 2021 and investigate a range of schedules of relaxation from lockdown. Prior to January 2021, the model assumes the same levels of control that took place in England. In this scenario, we introduce vaccination into the population at a rate of either 2 million or 4 million individuals per week. We assume a vaccine efficacy of 70% after the first dose and 88% after the second dose against disease (and 48% and 60% against infection), with immunity taking effect 2 weeks after vaccination. We investigate the optimal policy to emerge from lockdown and how that policy is dependent upon assumptions around the acceptable cost of control.

### Defining the counterfactual scenarios

In order to understand the health and economic impact of any level of control, for each scenario we must outline the counterfactual against which all other strategies are judged. For analytical purposes we compare all levels of control against an unmitigated outbreak (i.e. zero control, or *ϕ* = 0) without vaccination as our null model for the first scenario and a scenario where we stay in full lockdown (phi =) for the duration of the simulations, until the end of 2021. While this is unlikely in reality, as populations are likely to change behaviours as the risk of infection changes, it provides the base to assess different increasing levels of control (through either voluntary or mandatory distancing). In addition, throughout our analyses for simplicity we assumed that there are no economic losses as a result of reduced productivity from sickness and death. This means that any increase in the level of control always leads to an economic loss but a health benefit.

### Calculating costs and willingness to pay

For each simulation of the model, we calculated the total number of deaths by age and used this to establish a total QALY loss as a direct result of severe infection, taking into account mortality, hospitalisation and admission into intensive care (see Supporting Information). No discount rate was applied to future costs and QALYs given the limited timeframe of the analysis. We estimate economic loss as a result of a given set of potential control measures by investigating the total GDP loss related to the level of control. We approximated daily GDP by a polynomial function of the level of control, *ϕ* (Fig. 1, red line), derived from data on GDP from the period January to December 2020. Hence, we made a pessimistic assumption about the economic impact of control measures. Given the volatility in GDP in the months following the first national lockdown in March 2020, we make two assumptions, one in which we fit to the whole of 2020 (Fig. 1, red line) and one in which we fit from June 2020 to December 2020 (Fig. 1, blue line). We conjecture that the model fitted from June 2020 may be representative of the true long term impact of the pandemic and the underlying intervention measures upon GDP. We recognise using productivity as our measure of economic loss and using the blunt relationship between control and GDP, excludes the economic value of non-monetary elements of social welfare. Moreover, using an empirically derived relationship between control and GDP from the past, limits the policy inference, as observed levels of control are likely to be driven by both voluntary control (in response to levels of risk of illness) and mandatory lockdown measures.

**Fig. 1:**
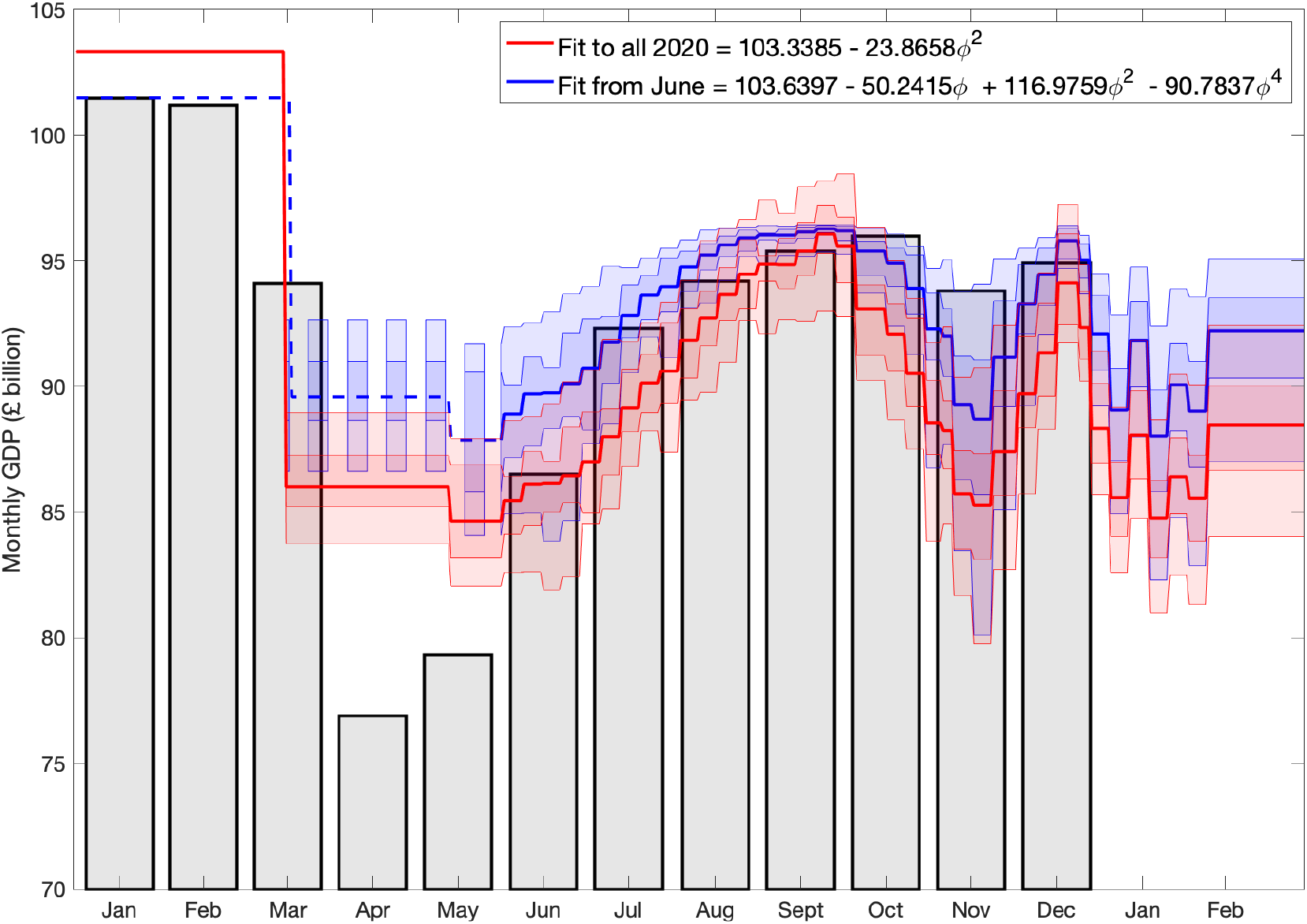
Graph of monthly GDP (in billions of pounds) from January to December 2020. Grey bars correspond to the GDP in the respective month. The red line shows the best (quadratic) fit to the GDP from January to December 2020, whilst the blue line shows the best (quartic) fit to the data from June 2020 to December 2020. (Shading regions show 95% credible intervals.)

To inform the temporal strength of controls, we used the temporally varying level of control inferred by the SARS-CoV-2 transmission model being fit to multiple epidemiological data streams as outlined in previous work [13]. Using our estimates of strength of controls, given a level of control at a particular point in time, we determined the polynomial function that provided the best fit to reported monthly GDP. We translated this value to total GDP loss by summing across the intervention duration and comparing to the unmitigated scenario.

To produce an optimal level of control from a societal perspective, we combined the value of health and economic loss estimates into a single monetary quantity. We achieved this by placing a monetary value (*W*) on each QALY (Quality Adjusted Life Year) and minimising the net monetary loss i.e. *W* × QALY loss + GDP loss. *W* represents the societal willingness to pay per QALY loss avoided. We model this for different levels of *W*. WTP is determined by decision authorities and may reflect different values. For example, the National Institute for Health and Care Excellence (NICE) currently uses a cost-effectiveness threshold where *W* is in the range of £20,000 to £30,000 for reimbursing new drugs in the National Health Service [16]; however, this is for policy decisions that considered trade-offs between new and current health sector services and in the context of the ongoing COVID-19 pandemic, we may expect a different value of *W* to be appropriate. We therefore consider a range of values of *W* from £20,000 to £200,000 throughout this paper.

## Results

### Analysis of the impact of precautionary breaks in 2020

We firstly investigated the effect that multiple precautionary break policies could have had on the epidemic from June 2020. For this set of results, we do not consider the effect of vaccination and we excluded simulations in which hospital occupancy across the entire country exceeded a specified threshold (set at either 10,000 or 20,000 individuals at any one time and we also include a scenario with no limit on hospital occupancy) to ensure that the optimal strategy did not result in the health service being overwhelmed. Whilst we imposed a health service related constraint here, our model framework can be extended to include other exclusionary constraints; for example, simulations that result in daily economic costs exceeding a given threshold or those that exceed specific hospitalisation thresholds by local area.

To exemplify the dynamics, we fixed the willingness to pay per QALY (*W*) at £50,000 and examined the net loss across a range of scenarios (Fig. 2) with no limit on hospital occupancy initially. As the level of control increases, the total QALY loss decreases, whilst the total GDP also decreases. We therefore observe that the net loss is minimised at an intermediate background level of control – a value of 0.7 for the given value of *W* when we fit GDP to all 2020 and 0.8 when we fit from June to December 2020. It is also optimal in this instance for two precautionary breaks to be introduced, in July/August 2020 and September 2020. We explored sensitivity to these findings by considering alternative fixed *W* values of £20,000, £30,000, £100,000 and £200,000. The results are summarised in Figures S1-S4. When considering £200,000 as a willingness to pay per QALY, we observe a slight increase in the optimal level of control, whilst if our willingness to pay per QALY is only £20,000, then the level of control that minimises loss is reduced.

**Fig. 2:**
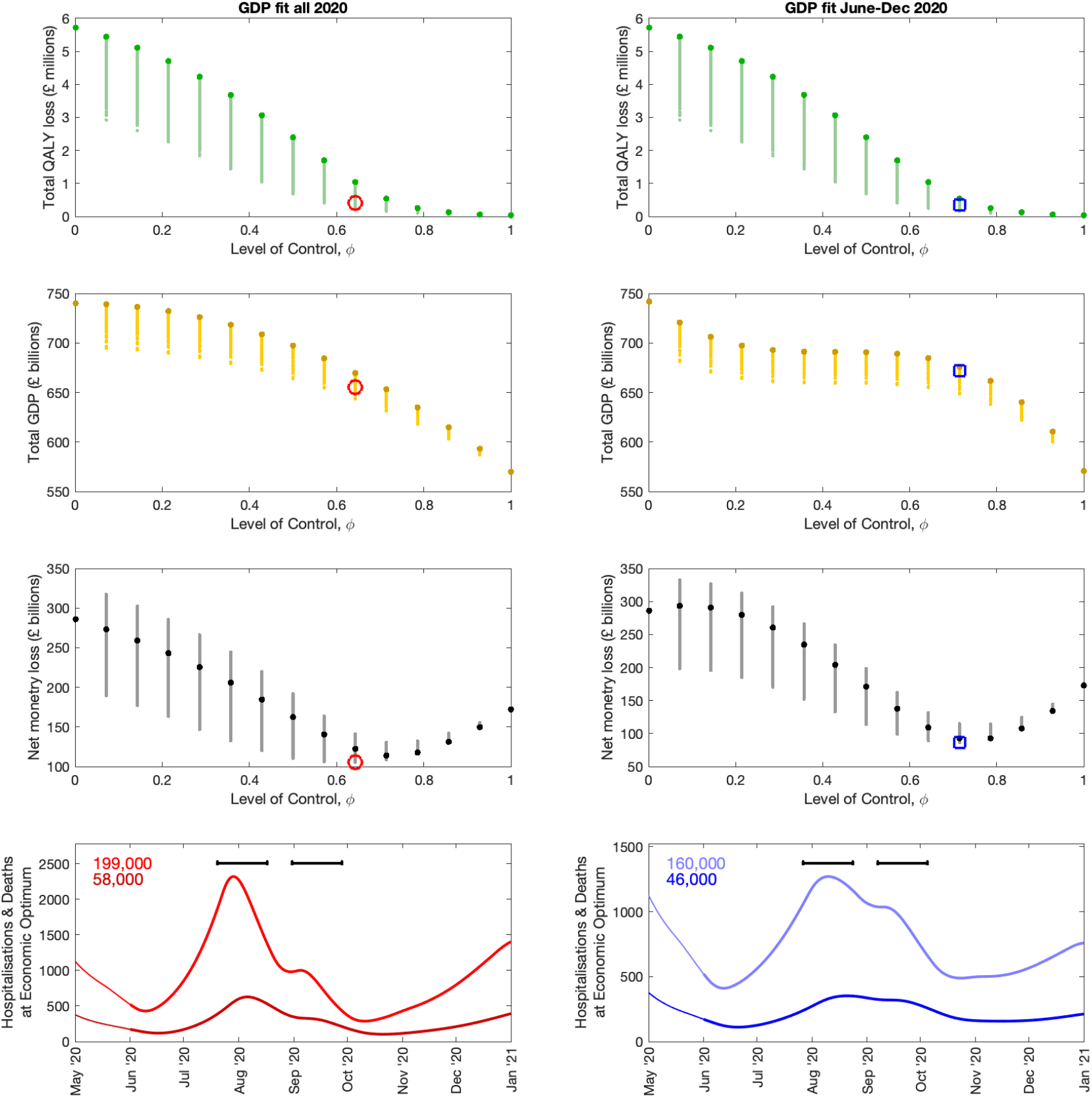
Outcomes at a willingness to pay per QALY (*W*) of £50,000 with no limit on hospital occupancy. For various levels of control, we present **(top row)** Total QALY loss, **(second row)** total GDP, **(third row)** net monetary loss (*W* × QALY loss + GDP loss) and **(fourth row)** hospitalisations and deaths. In these panels, darker, larger, dots are for a constant intrinsic control level without any planned short-term breaks, whilst the smaller dots represent different timings and frequency of precautionary breaks. The red circle (left column) and blue square (right column) indicate the minimum net monetary loss for *W* =£100,000 when there is no limit on daily hospitalisations. The bottom panels show, for the economic optimum, daily deaths (darker colour) and hospitalisations (lighter colour) with the total number of each given in the top left hand corner of each panel. The black bars represent when the precautionary breaks take place in each instance. In this figure, the left column shows the results when GDP is fitted to the whole of 2020, whilst the right column shows the results when GDP is fitted from June to December 2020.

We extended this approach across a range of *W* values, finding the strategy that minimised the net loss. Our results are summarised in Fig. 3 when we fit GDP from June to December 2020 (results when we fit to all 2020 are given in the supplementary material, **??**), with particular realisations at the optimal level of control shown in Figures S6-S11. We observe that, if the willingness to pay per QALY (*W*) was low and our restriction on daily hospital admissions was unlimited (blue curves), the optimal strategy was to have low levels of control outside the precautionary breaks and a relatively low control level within a precautionary break (Fig. 3 and **??**, second row, right panel and third row, right panel, respectively). As the willingness to pay increases, there is a preference for higher levels of control both within and outside the precautionary break and a preference for a slightly delayed introduction of the second precautionary break, but regardless of the willingness to pay, the tendency is for both the first and second precautionary break to be introduced early. We note that for almost all scenarios investigated, the optimal duration of a precautionary break was the maximum length investigated of 4 weeks (Fig. 3 and **??**, third row, right hand panel), whilst multiple rather than single breaks were generally preferred. This suggests that there may be a need for either longer duration or more frequent breaks to be investigated. We also note that the limit on hospital occupancy only has a significant effect upon the optimal policy when the willingness to pay is low - at higher values willingness to pay the preferred timing and intensity of the precautionary break is found to be independent of the hospital occupancy threshold.

**Fig. 3:**
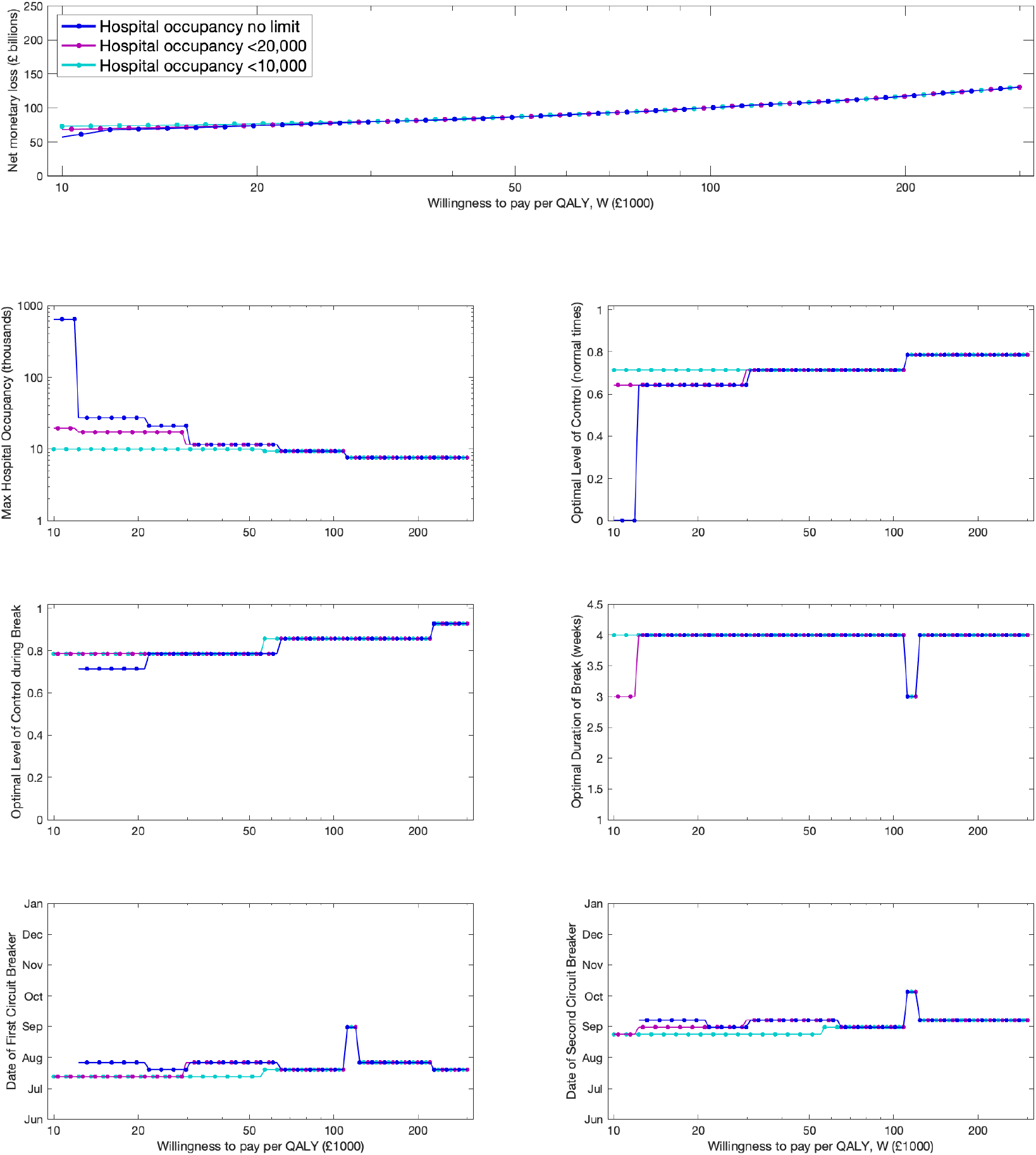
Outcomes over a range of willingness to pay per QALY (*W*) values. (Top Row)Net monetary loss (W × QALY loss + GDP loss) against different values of *W* as the daily hospitalisation threshold varies (different colours). In rows two-four, we display the following measures for the optimal control strategy as the willingness to pay per QALY increases: **(second row, left panel)** maximum number of hospital admissions per day. **(second row, right panel)** the optimal level of intrinsic control outside lockdown; **(third row, left panel)** the optimal level of control within a precautionary break; **(third row, right panel)** the optimal duration of the lock-down in days; **(fourth row, left panel)** the optimal date of the first precautionary break; **(fourth row, right panel)** the optimal date of the second precautionary break. In this figure we fit to GDP from June to December 2020.

In the supplementary material, we used the same three hospitalisation thresholds and show the dynamics of deaths and hospitalisations at the optimal set of controls that minimises the net monetary loss for particular values of *W* (*W* =£20,000, £30,000, £50,000, £100,000 and £200,000). Across all willingness to pay values, we find that the optimal policy is for two precautionary breaks to be introduced early, with a lower value of willingness to pay generally resulting in a slightly early date of introduction for the precautionary breaks. As the willingness to pay increases, the optimal intensity of control both within and outside the precautionary break periods increases and results in fewer daily hospitalisations and deaths (Figures S6-S11). We also note that increasing the maximum number of hospitalisations per day threshold does not significantly affect the optimal timing of precautionary breaks but allows for lower background levels of control.

### Optimal relaxation strategies for the January 2021 national lockdown

We now investigate how the optimal pace of relaxation of lockdown may be dependent upon our societal willingness to pay for repeated lockdowns to avoid QALY losses due to COVID. The results are summarised in Fig. 4. Here we can see that, if the willingness to pay is at the low end of our range and 4 million individuals are vaccinated per week (dark colours in figures), our model recommends a relatively rapid relaxation policy, with all interventions removed by September 2021. However, this results in a significant wave of deaths occurring. As the willingness to pay increases, it is preferable to ease out of lockdown much more gradually, as this results in a much lower peak in deaths as non-pharmaceutical interventions are relaxed. If we only have the capacity to vaccinate 2 million individuals per week, we note that this results in a slightly slower easing of lockdown being optimal and as a result of this, we observe a reduced peak in deaths for a given willingness to pay when compared with a scenario with a higher vaccine capacity. These results highlight that the willingness to pay has a significant influence upon the optimal strategy for both introducing and easing interventions.

**Fig. 4:**
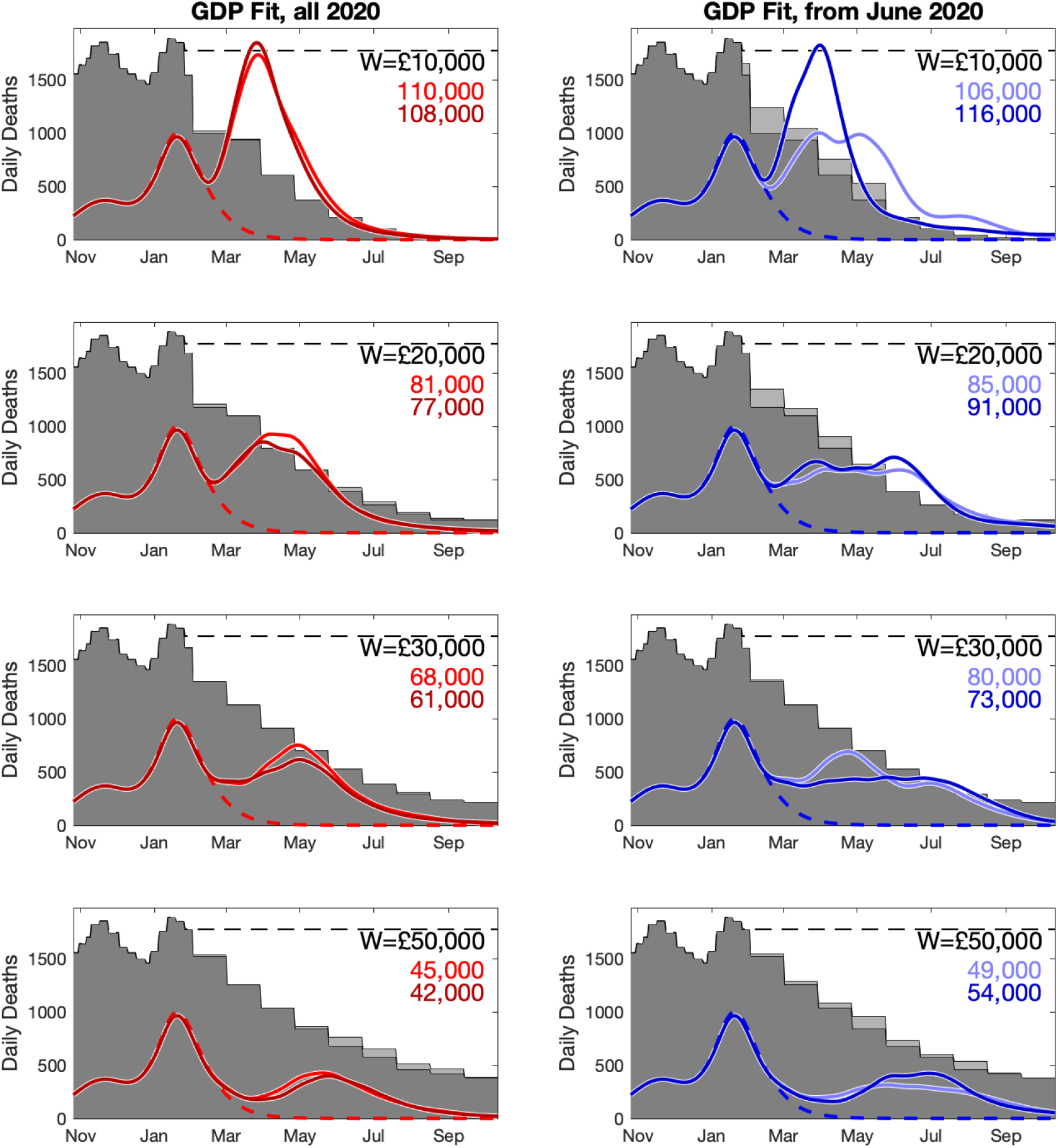
Optimal relaxation policies from the January 2021 lockdown over a range of willingness to pay per QALY (*W*) values. Daily deaths from November 2020 to September 2021 when 2 million individuals (dark colours) and 4 million individuals (light colours) are vaccinated per day. The background grey shaded region shows the relative level of control required at the optimal value whilst the dashed line shows the daily deaths when full lockdown is in place throughout. The left column shows results when we fit to GDP for the whole of 2020 whilst the right column is for when we fit from June to December 2020. Each row shows results for a different value of willingness to pay, from £10,000 to £50,000 per QALY gained.

## Discussion

This work highlights a central tenet of disease control - that the optimal timing and level of any physical distancing policy is highly dependent upon the scope of the objective that is trying to be achieved [17, 18]. Combining economic and health outcomes in COVID-19 policy analysis using infectious disease modelling is potentially complex. Our methods here are a starting point, but nevertheless illustrate that where a multi-phase precautionary break strategy is under consideration, then the exact timescale and intensity of such a policy is highly dependent upon the willingness to pay of the UK population to averted health loss. For example, in our retrospective scenario, if WTP is £50,000 per QALY loss avoided, increasing the level of control from 0 to 0.7 would result in a reduction in net monetary loss of over £150 billion. If the WTP is £100,000 then this change in control level would result in a increased reduction in net monetary loss to over £400 billion. Even where the exact WTP is unknown, our analysis illustrates that it is possible to develop an approach for arriving at optimal control that incorporates both health and economic losses, for different levels of willingness to pay, with the aim to inform the transparent assessment of the trade-offs involved.

There are important caveats to the findings in this work that should be discussed, and can inform those who wish to develop this approach further. First, we have examined the relationship between levels of control and economic and health impact. The level of control is driven both by voluntary and mandated distancing, and the former is likely to be driven at least in part by economic and health risk. These feed backs may result in underestimating the optimal level of control, since strong controls can reduce disease incidence and hence minimise economic damage from voluntary distancing. We also note that our economic approach, utilising current GDP loss as a proxy for overall economic loss, is unlikely to capture the dynamics of economic impact and wider societal welfare and long-term economic impacts of control, particularly given government borrowing to mitigate costs in the short term. There will be a range of lagged impacts, and longer-term impacts on GDP of any set of controls. We also note that GDP loss is not the only important economic indicator. Fiscal impact or increases in unemployment are other important factors, and wider impacts such as educational losses could be considered. It is therefore possible that the findings represented here may overestimate the short-term economic impact of lockdown, whilst not capturing the social and long-term economic impact. These aspects should also be considered when determining the optimal strength of control implemented within lockdown periods. A more complete economic analysis would use approaches such as computable general equilibrium models [19] and the need to consider which sectors of society would be restricted to arrive at a particular level of control, and how this would lead to a loss in GDP. Our findings do suggest, however, that governments should consider setting acceptable thresholds for a loss of productivity per QALY before the next pandemic in order to establish optimal intervention policies at pandemic onset.

On measuring cost to health, we currently estimate health harms by calculating QALY loss from hos-pitalisation, admission to intensive care and death as a result of COVID-19 infection. However, a more complete analysis would account for a range of other factors beyond the scope of this preliminary work, including: non-COVID health impacts of COVID-19 itself and of interventions to control COVID-19 (both positive, such as the likely decline in other infections due to social distancing, and negative, such as excess all-cause mortality, cancelled elective procedures due to hospital bed pressures, and reduction in hospital attendance and screening), the mental health and social impacts of COVID-19 and its control [20], long-term health impacts of economic harm [21] and the lasting health implications of long-COVID [22, 23].

We also note that our results are highly dependent upon the precise value of the Willingness to Pay, *W*. When faced with the scale of the SARS-CoV-2 outbreak and associated COVID-19 disease burden, trade-offs may consider a rule of rescue and the value of *W* could be higher than values typically used by the NHS [24]. Alternative approaches, using measures of *W* that value health according to the opportunity costs within current spending on health are likely to be lower. Finally, depending on the way in which *W* is determined, there may be some double counting between economic loss and health loss as, when valuing QALYs, the value of reduced productivity may be implicitly considered in QALY valuations. Given this complexity around valuing health improvement, we present our results for a range of different values of *W*.

It is also important to note that our models assumed a fixed rate of adherence to intervention measures, a fixed level of control across all lockdown periods and similarly a fixed (but lower) level of control across non-lockdown periods. It is possible that such a policy of repeated lockdowns could result in waning adherence over time, and this may in turn influence the overall economic and health impact of any lockdown period. With this in mind, further work may be needed in order to establish the optimal adaptive policy when there is uncertainty regarding future intensities of lockdown and adherence to intervention measures. Finally, economic cost is proportional to the total duration and intensity of a lockdown. We have not considered any additional economic cost of multiple short precautionary breaks (owing to stopping and starting of businesses) or longer term lockdowns (in which businesses may be more likely to become insolvent).

Despite the caveats presented above, there are some important lessons that we can learn from this research. During any infectious disease outbreak, policy makers must rapidly evaluate the state of the system and introduce an intervention policy that is deemed most appropriate at that point in time. For human diseases, the key focus is typically (and understandably) on health, with policies often selected that will minimise the risk of severe disease or death over a short period of time. However, a policy that focuses purely upon minimising health losses can have a very high macro-economic cost and result in long-term harm. Here, we present a more nuanced approach, whereby we consider both the economic costs of lockdown and the health costs of the pandemic, linked to a dynamic model of infectious disease. Our results highlight the necessity for decision makers to identify their overarching objective in the event of an infectious disease outbreak. We concede it may be challenging to do this during a pandemic when there are multiple competing priorities facing decision makers, and population preferences are unclear. However, clearly defining the objective and the trade offs that need to be considered should be an essential component of the contingency planning process going forward, and can help ensure that optimal policies can be defined to minimise the impact of a future epidemic rapidly. Once an appropriate objective is decided upon, the research presented here provides the start of a framework for decision makers to evaluate the effectiveness from a societal perspective.

## Data Availability

Data on cases were obtained from the COVID-19 Hospitalisation in England Surveillance System (CHESS) data set that collects detailed data on patients infected with COVID-19. Data on COVID-19 deaths were obtained from Public Health England. These data contain confidential information, with public data deposition non-permissible for socioeconomic reasons. The CHESS data resides with the National Health Service (www.nhs.gov.uk) whilst the death data are available from Public Health England (www.phe.gov.uk).

## Author contributions

**Conceptualisation:** Michael J. Tildesley; Matt J. Keeling; Steven Riley.

**Data curation:** Matt J. Keeling.

**Formal analysis:** Matt J. Keeling; Michael J. Tildesley; Steven Riley; Anna Vassall; Mark Jit; Frank Sandmann.

**Investigation:** Michael J. Tildesley; Matt J. Keeling; Anna Vassall.

**Methodology:** Matt J. Keeling; Michael J. Tildesley; Anna Vassall; Steven Riley; Mark Jit; Frank Sandmann.

**Software:** Matt J. Keeling.

**Validation:** Matt J. Keeling; Edward M. Hill; Louise Dyson; Michael J. Tildesley; Benjamin D. Atkins; Robin N. Thompson; Anna Vassall; John Edmunds; Frank Sandmann; Mark Jit; Steven Riley.

**Visualisation:** Matt J. Keeling.

**Writing - original draft:** Michael J. Tildesley; Anna Vassall.

**Writing - review & editing:** Mark Jit; Anna Vassall; Frank Sandmann; Matt J. Keeling; Edward M. Hill; Louise Dyson; Robin N. Thompson; Benjamin D. Atkins; John Edmunds; Steven Riley; Michael J. Tildesley.

## Financial disclosure

This work has been supported by the Engineering and Physical Sciences Research Council through the MathSys CDT [grant number EP/S022244/1] and by the Medical Research Council through the COVID-19 Rapid Response Rolling Call [grant number MR/V009761/1]. MJ and WJE were supported by the NIHR Heath Protection Research Units in Immunisation (NIHR200929) and Modelling and Health Economics (NIHR200908), as well as by the European Commission project Epipose (101003688). The funders had no role in study design, data collection and analysis, decision to publish, or preparation of the manuscript.

## Data availability

## Competing interests

All authors declare that they have no competing interests.

## Notes

### Competing Interest Statement

The authors have declared no competing interest.

### Clinical Trial

N/A

### Author Declarations

The data were supplied from the CHESS database after anonymisation under strict data protection protocols agreed between the University of Warwick and Public Health England. The ethics of the use of these data for these purposes was agreed by Public Health England with the Governments SPI-M(O) / SAGE committees.

