## Supplementary Information for "Optimal health and economic impact of non-pharmaceutical intervention measures prior and post vaccination in England: a mathematical modelling study"

### 1 Model description

Here we present the system of equations that account for both household saturation of transmission and household quarantining. Individuals may be susceptible ($S$), exposed ($E$), with detectable infection ($D$), or undetectable infection (asymptomatic, $U$). Undetectable infections are assumed to transmit infection at a reduced rate given by $\tau$. We let superscripts denote the first infection in a household ($F$), a subsequent infection from a detectable/symptomatic household member ($SD$) and a subsequent infection from an asymptomatic household member ($SU$). A fraction ($H$) of the first detected case in a household is quarantined ($QF$), as are all their subsequent household infections ($QS$).

#### Model equations

The full equations are given by

$$
\begin{aligned}
\frac{dS_a}{dt} &= -\left(\lambda_a^F + \lambda_a^{SD} + \lambda_a^{SU} + \lambda_a^Q\right)\frac{S_a}{N_a}, \\
\frac{dE_a^F}{dt} &= \lambda_a^F \frac{S_a}{N_a} - \epsilon E_a^F, \\
\frac{dE_a^{SD}}{dt} &= \lambda^{SD} \frac{S_a}{N_a} - \epsilon E_a^{SD}, \\
\frac{dE_a^{SU}}{dt} &= \lambda^{SU} \frac{S_a}{N_a} - \epsilon E_a^{SU}, \\
\frac{dE_a^Q}{dt} &= \lambda^Q S - \epsilon E_a^Q, \\
\frac{dD_a^F}{dt} &= d_a(1-H)\epsilon E_a^F - \gamma D_a^F, \\
\frac{dD_a^{SD}}{dt} &= d_a \epsilon E_a^{SD} - \gamma D_a^{SD}, \\
\frac{dD_a^{SU}}{dt} &= d_a(1-H)\epsilon E_a^{SU} - \gamma D_a^{SU}, \\
\frac{dD_a^{QF}}{dt} &= d_a H \epsilon E_a^F - \gamma D_a^{QF}, \\
\frac{dD_a^{QS}}{dt} &= d_a H \epsilon E_a^{SU} + d_a \epsilon E_a^Q - \gamma D_a^{QS}, \\
\frac{dU_a^F}{dt} &= (1-d_a)\epsilon E_a^F - \gamma U_a^F, \\
\frac{dU_a^S}{dt} &= (1-d_a)\epsilon (E_a^{SD} + E_a^{SU}) - \gamma U_a^S, \\
\frac{dU_a^Q}{dt} &= (1-d_a)\epsilon E_a^Q - \gamma U_a^Q,
\end{aligned}
$$

with the forces of infection obeying

$$\begin{aligned}
\lambda_a^F &= \sigma_a \sum_b \left( D_b^F + D_b^{SD} + D_b^{SU} + \tau (U_b^F + U_b^S) \right) \beta_{ba}^N, \\
\lambda_a^{SD} &= \sigma_a \sum_b D_b^F \beta_{ba}^H, \\
\lambda_a^{SU} &= \sigma_a \tau \sum_b U_a^F \beta_{ba}^H, \\
\lambda_a^Q &= \sigma_a \sum_b D_b^{QF} \beta_{ba}^H,
\end{aligned}$$

where $\beta_{ba}^H$ (with the subscript $ba$ corresponding to transmission from age group $b$ towards age group $a$) is household transmission and $\beta_{ba}^N = \beta_{ba}^S + \beta_{ba}^W + \beta_{ba}^O$ is all other transmission locations, comprising school-based transmission ($\beta_{ba}^S$), work-place transmission ($\beta_{ba}^W$) and transmission in all other locations ($\beta_{ba}^O$). $\sigma_a$ corresponds to the age-dependent susceptibility of individuals to infection, $d_a$ the age-dependent probability of displaying symptoms (and hence being detected), and $\tau$ represents reduced transmission of infection by undetectable individuals compared to detectable infections.

##### Relationship between age-dependent susceptibility and detectability

We interlink age-dependent susceptibility, $\sigma_a$, and detectability, $d_a$, by a quantity $Q_a$. $Q_a$ can be viewed as the scaling between force of infection and symptomatic infection. Taking a next-generation approach, the early dynamics would be specified by:

$$R_0 D_a = d_a \sigma_a \beta_{ba}^N \left( D_a + \tau U_a \right) / \gamma \qquad R_0 U_a = (1 - d_a) \sigma_a \beta_{ba}^N \left( D_a + \tau U_a \right) / \gamma$$

where $D_a$ measures those with detectable infections, which mirrors the early recorded age distribution of symptomatic cases. Explicitly, we let $d_a = \frac{1}{\kappa} Q_a^{(1-\alpha)}$ and $\sigma_a = \frac{1}{k} Q_a^{\alpha}$. As a consequence, $Q_a = \kappa k d_a \sigma_a$; where the parameters $\kappa$ and $k$ are determined such that the oldest age groups have a 90% probability of being symptomatic ($d_{>90} = 0.90$) and such that the basic reproductive ratio from these calculations gives $R_0 = 2.7$.

#### 2 Public health measurable quantities

In order to be able to interpret our model and compare the outputs to the data, we considered five quantities which we calculated from the number of newly detectable symptomatic infections on a given day $nD_d$.

1. **Hospital Admissions:** We assume that a fraction $P_a^{D\to H}$ of detectable cases will be admitted into hospital after a delay $q$ from the onset of symptoms. The delay, $q$, is drawn from a distribution $D_q^{D\to H}$ (note that $\sum_q D_q^{D\to H} = 1$.) Hospital admissions on day $d$ of age $a$ are therefore given by
$$H_a(d) = P_a^{D\to H} \sum_q D_q^{D\to H} nD_{d-q}$$

2. **ICU Admissions:** Similarly, a fraction $P_a^{D\to I}$ of detectable cases will be admitted into ICU after a delay, drawn from a distribution $D_q^{D\to I}$ which determines the time between the onset of symptoms and admission to ICU. ICU admissions on day $d$ of age $a$ are therefore given by
$$ICU_a(d) = P_a^{D\to I} \sum_q D_q^{D\to I} nD_{d-q}$$

3. **Hospital Beds Occupied:** Individuals admitted to hospital spend a variable number of days in hospital. We therefore define two weightings, which determine if someone admitted to hospital still occupies a hospital bed $q$ days later ($T_q^H$) and if someone admitted to ICU occupies a hospital bed on a normal ward $q$ days later ($T_q^{I\to H}$). Hospital beds occupied on day $d$ of age $a$ are therefore given by
$$H_a^o(d) = \sum_q H_a(d-q) T_q^H + \sum_q ICU_a(d-q) T_q^{I\to H}$$

4. **ICU Beds Occupied:** We similarly define $T_q^I$ as the probability that someone admitted to ICU is still occupying a bed in ICU $q$ days later. ICU beds occupied on day $d$ of age $a$ are therefore given by
$$ICU_a^o(d) = \sum_q ICU_a(d-q) T_q^I$$

5. **Number of Deaths:** The mortality ratio $P_a^{H\to Death}$ determines the probability that a hospitalised case of a given age, $a$, dies after a delay, $q$ drawn from a distribution, $D_d^{H\to Death}$ between hospitalisation and death. The number of deaths on day $d$ of age $a$ are therefore given by
$$\text{Deaths}_a(d) = P_a^{H\to Death} \sum_q H_a(d-q) D_d^{H\to Death}$$

These nine distributions are all parameterised from individual patient data as recorded by the COVID-19 Hospitalisation in England Surveillance System (CHESS) [1].

However, these distributions all represent a national average and do not therefore reflect regional differences. We therefore define regional scalings of the three key probabilities ($P_a^{D\to H}$, $P_a^{D\to I}$ and $P_a^{H\to Death}$) and two additional parameters that can stretch (or contract) the distribution of times spent in hospital and ICU. These five regional parameters are necessary to get good agreement between key observations in all regions and may reflect both differences in risk groups (in addition to age) between regions or differences in how the data are recorded between devolved nations. We stress that these parameters do not (of themselves) influence the epidemiological dynamics, but do strongly influence how we fit to the evolving dynamics.

#### 3 Modelling social distancing

Age-structured contact matrices for the United Kingdom were obtained from Prem et al. [2] and used to provide information on household transmission ($\beta^H_{ba}$, with the subscript $ba$ corresponding to transmission from age group $b$ towards age group $a$), school-based transmission ($\beta^S_{ba}$), work-place transmission ($\beta^W_{ba}$) and transmission in all other locations ($\beta^O_{ba}$). We assumed that the suite of social-distancing and lockdown measured acted in concert to reduce the work, school and other matrices while increasing the strength of household contacts.

We capture the impact of physical distancing by defining new transmission matrices ($B_{ba}$) that represent the potential transmission in the presence of extreme lockdown. In particular, we assume that:

$$B^S_{ba} = q^S \beta^S_{ba}, \qquad B^W_{ba} = q^W \beta^W_{ba}, \qquad B^O_{ba} = q^O \beta^O_{ba},$$

while household mixing $B^H$ is increased by up to a quarter to account for the greater time spent at home. We take $q^S = 0.05$, $q^W = 0.2$ and $q^O = 0.05$ to approximate the reduction in attendance at school, attendance at workplaces and engagement with shopping and leisure activities during the lock-down, respectively.

For a given compliance level, $\phi$, we generate new transmission matrices as follows:

$$\begin{aligned}
\widehat{\beta}^H_{ba} &= (1-\phi)\beta^H_{ba} + \phi B^H_{ba} \\
\widehat{\beta}^S_{ba} &= (1-\phi)\beta^S_{ba} + \phi B^S_{ba} \\
\widehat{\beta}^W_{ba} &= (1-\theta)\left[(1-\phi)\beta^W_{ba} + \phi B^W_{ba}\right] + \theta\left((1-\phi) + \phi q^W\right)((1-\phi) + \phi q^O)\beta^W_{ba} \\
\widehat{\beta}^O_{ba} &= \beta^O_{ba}((1-\phi) + \phi q^O)^2
\end{aligned}$$

As such, home and school interactions are scaled between their pre-lockdown values ($\beta$) and post-lockdown limits ($B$) by the scaling parameter $\phi$. Work interactions that are not in public-facing 'industries' (a proportion 1 - $\theta$) were also assumed to scale in this manner; while those that interact with the general populations (such as shop-workers) were assumed to scale as both a function of their reduction and the reduction of others. We have assumed $\theta = 0.3$ throughout. Similarly, the reduction in transmission in other settings (generally shopping and leisure) has been assumed to scale with the reduction in activity of both members of any interaction, giving rise to a squared term.

### 4 QALY losses

Our computation of loss of quality adjusted life years (QALYs) incorporated loss due to deaths and losses associated with severe cases requiring hospitalisation.

QALY losses due to death were based on the quality adjusted life expectancy by age, modified for the relative life-expectancy of individuals that die:

$$\text{Fatal case QALY loss} = \sum_{a=1}^{21} \left(D(a) \times E(a)\right),$$

where $D(a)$ is the number of deaths in age bracket $a$, and $E(a)$ is the discounted quality adjusted value of the remaining life expectancy, $L(a)$, of individuals in age group $a$. This quality adjusted life expectancy is given by:

$$E(a) = \sum_{i=1}^{L(a)} \frac{Q_w(\hat{a} + i)}{(1 + d)^i}$$

where $Q_w(a)$ is the age-specific quality of life weight at age $a$, $\hat{a}$ is the average age (in years) of an individual in age-group $a$, and $d$ the discount rate (set at 0.035, corresponding to 3.5% per annum); the values of $L(a)$ are rounded to full years.

For individuals that are admitted to hospital (or ICU) we make the pessimistic assumption that their quality of life while in hospital is zero:

$$\text{Hospitalised QALY loss} = \sum_{a=1}^{21} \left(H(a) \times Q_w(a) \times \overline{H_S}\right),$$

where $H(a)$ is the number of hospital admissions in age bracket $a$, and $\overline{H_S}$ is the average hospital stay (approximately 10 days). We ignore the impact of recovery time outside the hospital and the effects of long-COVID. In all our calculations QALY loss from mortality vastly outweighs loss from hospital admissions.

For parameterising the age-specific quality of life weights, $Q_w$, we obtained age-specific EQ-5D index population norms estimates for England from two literature sources. We took childhood estimates (which we used to cover 0–19 years of age) from Table 3 of [3], and values for those aged 20 and above were sourced from Table 3.6 of [4]. A complete listing of age-specific quality of life weights values by age is presented in Table S1.

**Table S1: EQ-5D index population norms for England.**

| Age group (yrs) | EQ-5D index population norms scale |
|---|---|
| <20 | 0.948 |
| 20–24 | 0.929 |
| 25–34 | 0.919 |
| 35–44 | 0.893 |
| 45–54 | 0.855 |
| 55–64 | 0.810 |
| 65–74 | 0.773 |
| 75+ | 0.703 |

### 5 Additional figures

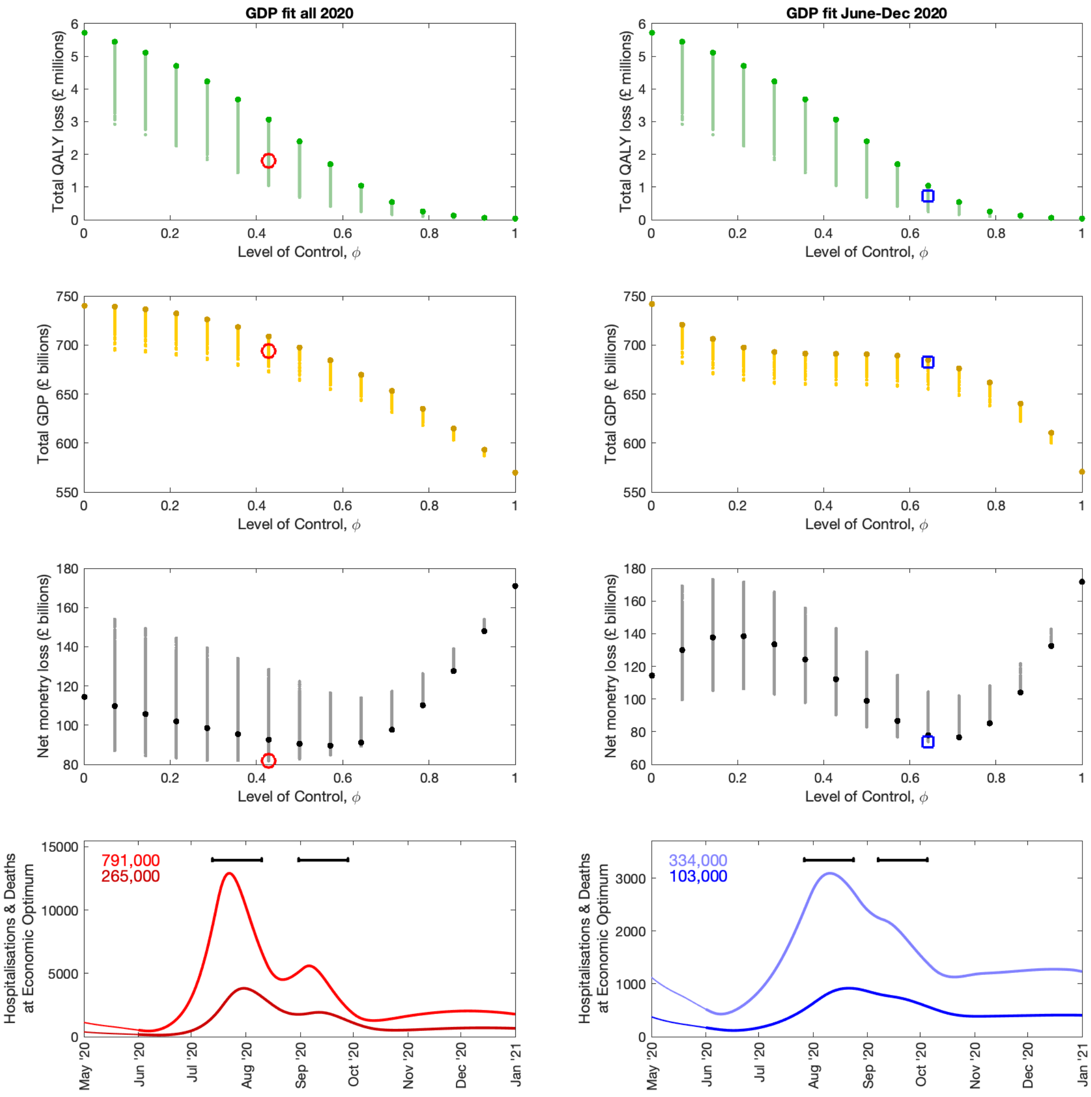


**Fig S1: Outcomes at a willingness to pay per QALY ($W$) of £20,000 with no limit on hospital occupancy.** For various levels of control, we present **(top row)** Total QALY loss, **(second row)** total GDP and **third row)** net monetary loss ($W$ × QALY loss + GDP loss). In these panels, darker,larger, dots are for a constant intrinsic control level without any planned short-term breaks, whilst the lighter dots represent different timings and frequency of precautionary breaks. The red circle (left column) and blue square (right column) indicate the minimum net monetary loss for $W$=£20,000 when there is no limit on daily hospitalisations. The bottom panels show, for the economic optimum, daily deaths (darker colour) and hospitalisations (lighter colour) with the total number of each given in the top left hand corner of each panel. The black bars represent when the precautionary breaks take place in each instance. In this figure, the left column shows the results when GDP is fitted to the whole of 2020, whilst the right column shows the results when GDP is fitted from June to December 2020.

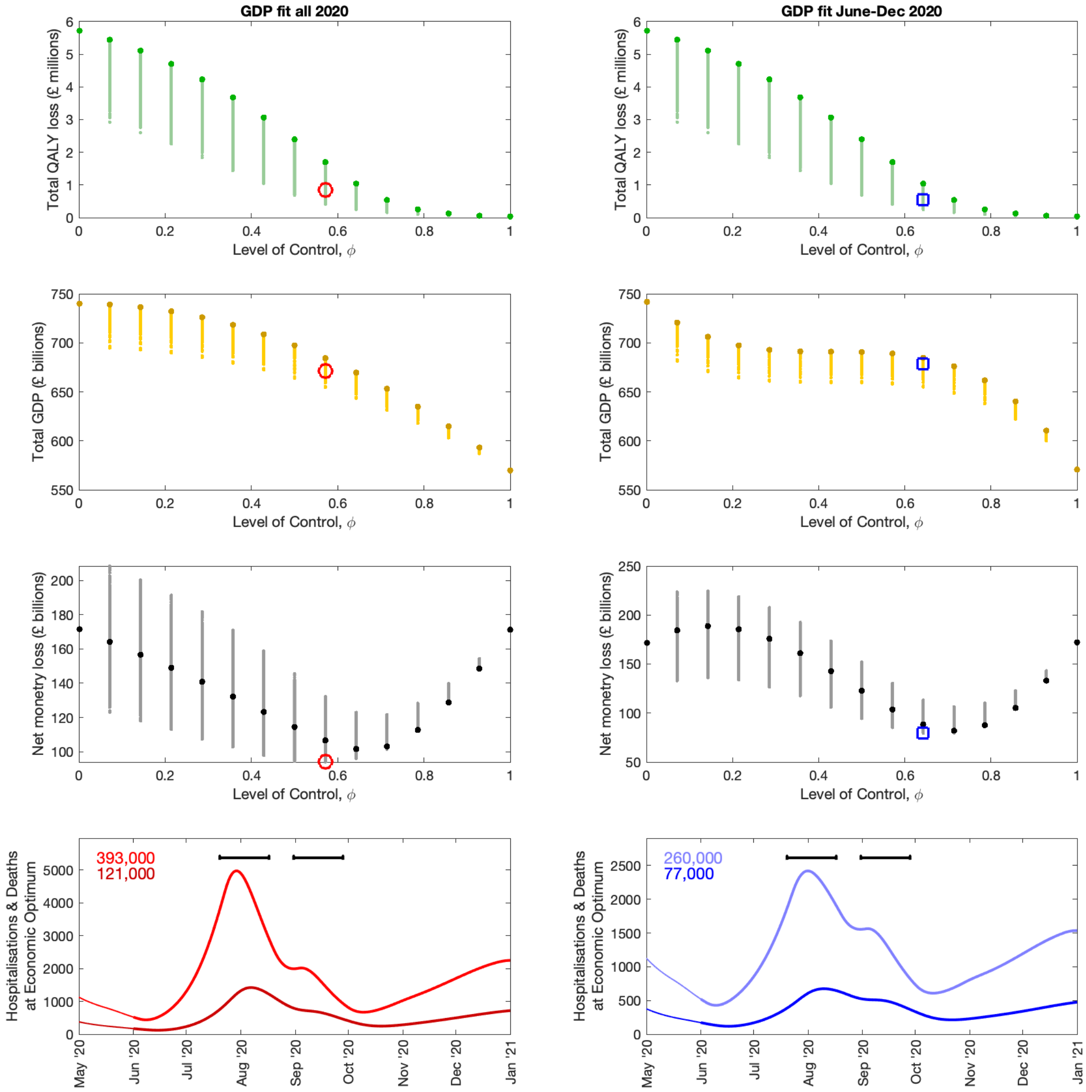


**Fig S2: Outcomes at a willingness to pay per QALY ($W$) of £30,000 with no limit on hospital occupancy.** For various levels of control, we present **(top row)** Total QALY loss, **(second row)** total GDP and **third row)** net monetary loss ($W \times$ QALY loss + GDP loss). In these panels, darker, larger, dots are for a constant intrinsic control level without any planned short-term breaks, whilst the lighter dots represent different timings and frequency of precautionary breaks. The red circle (left column) and blue square (right column) indicate the minimum net monetary loss for $W$=£30,000 when there is no limit on daily hospitalisations. The bottom panels show, for the economic optimum, daily deaths (darker colour) and hospitalisations (lighter colour) with the total number of each given in the top left hand corner of each panel. The black bars represent when the precautionary breaks take place in each instance. In this figure, the left column shows the results when GDP is fitted to the whole of 2020, whilst the right column shows the results when GDP is fitted from June to December 2020.

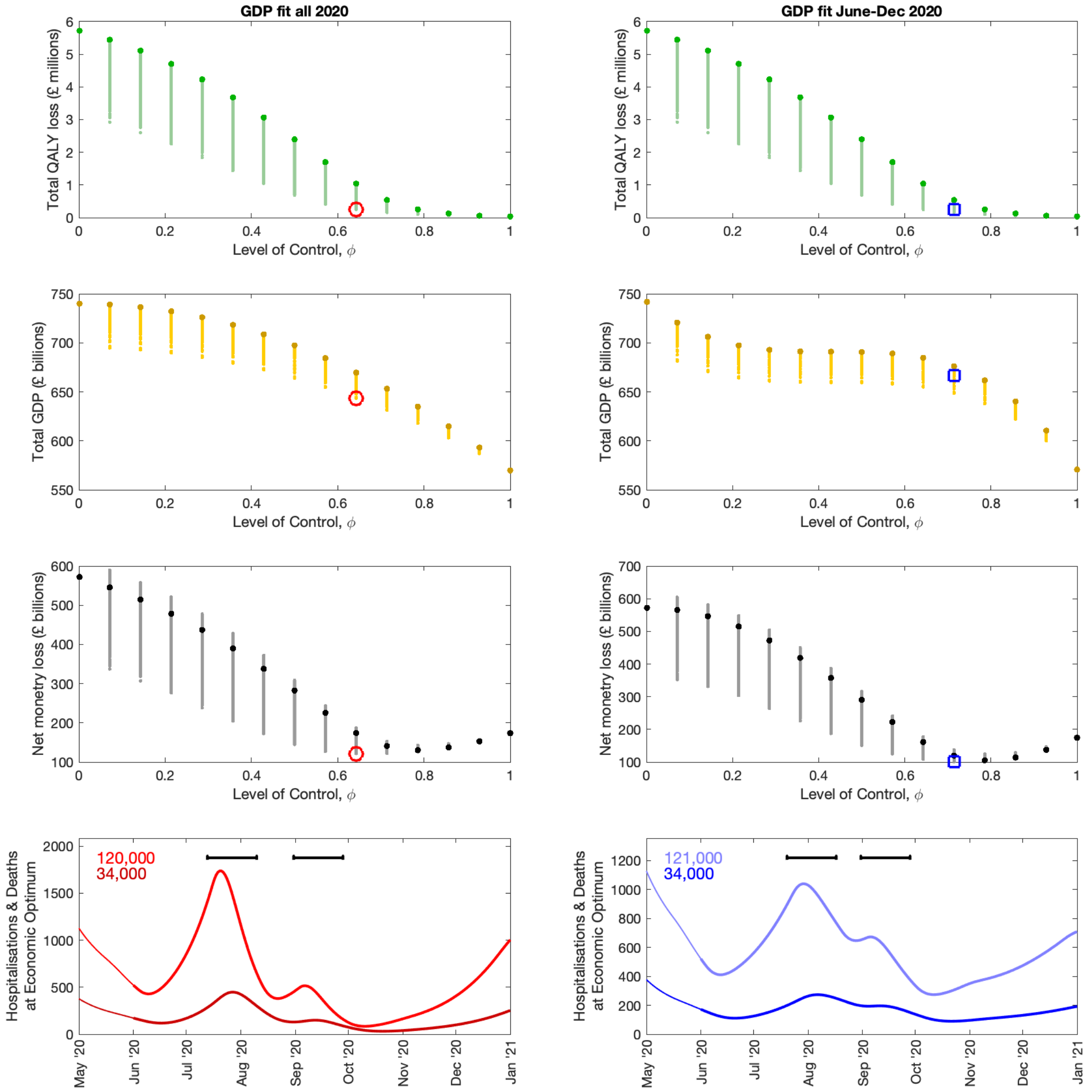


**Fig S3: Outcomes at a willingness to pay per QALY ($W$) of £100,000 with no limit on hospital occupancy.** For various levels of control, we present **(top row)** Total QALY loss, **(second row)** total GDP and **third row)** net monetary loss ($W \times$ QALY loss + GDP loss). In these panels, darker, larger, dots are for a constant intrinsic control level without any planned short-term breaks, whilst the lighter dots represent different timings and frequency of precautionary breaks. The red circle (left column) and blue square (right column) indicate the minimum net monetary loss for $W$=£100,000 when there is no limit on daily hospitalisations. The bottom panels show, for the economic optimum, daily deaths (darker colour) and hospitalisations (lighter colour) with the total number of each given in the top left hand corner of each panel. The black bars represent when the precautionary breaks take place in each instance. In this figure, the left column shows the results when GDP is fitted to the whole of 2020, whilst the right column shows the results when GDP is fitted from June to December 2020.

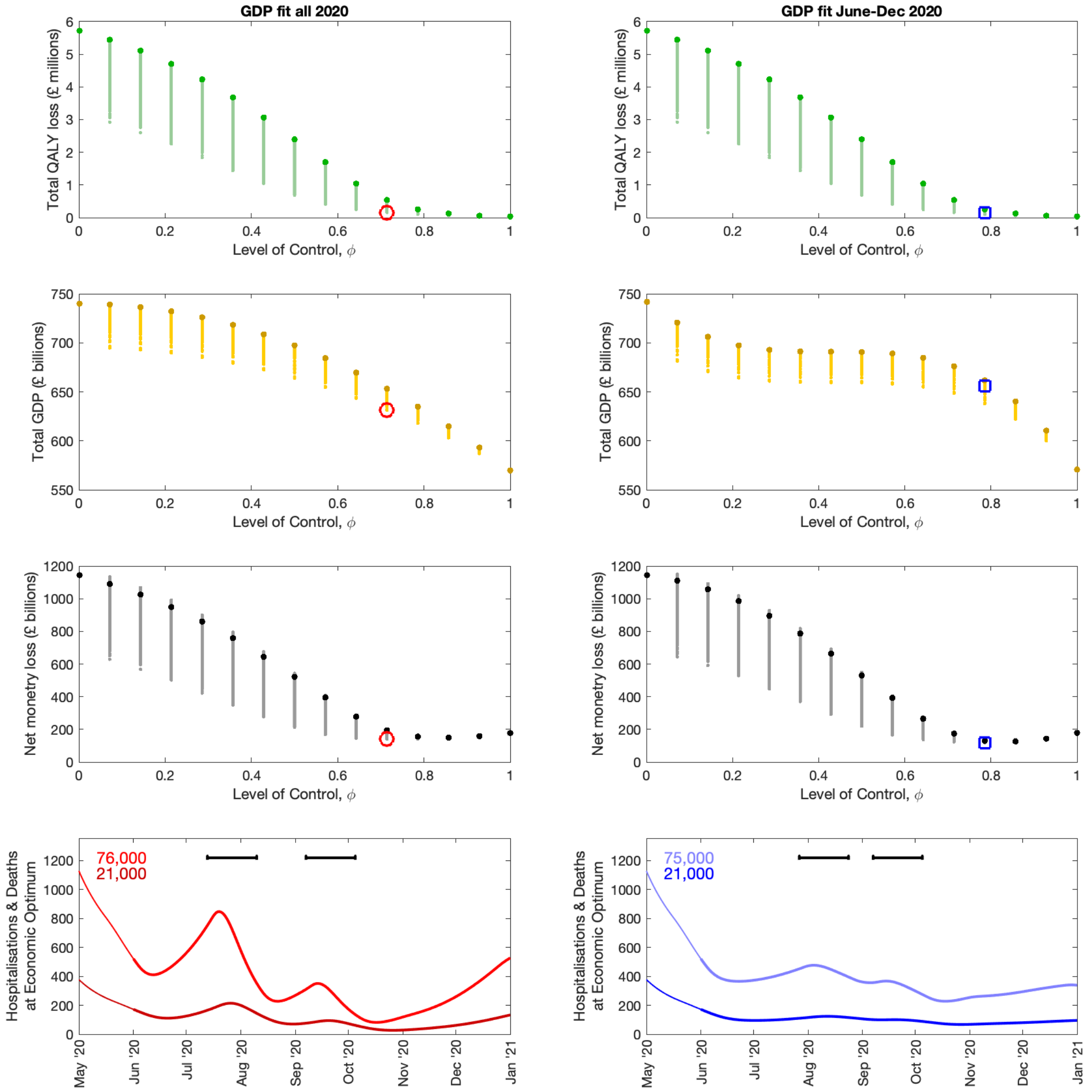


**Fig S4: Outcomes at a willingness to pay per QALY ($W$) of £200,000 with no limit on hospital occupancy.** For various levels of control, we present **(top row)** Total QALY loss, **(second row)** total GDP and **third row)** net monetary loss ($W$ × QALY loss + GDP loss). In these panels, darker, larger, dots are for a constant intrinsic control level without any planned short-term breaks, whilst the lighter dots represent different timings and frequency of precautionary breaks. The red circle (left column) and blue square (right column) indicate the minimum net monetary loss for $W$=£200,000 when there is no limit on daily hospitalisations. The bottom panels show, for the economic optimum, daily deaths (darker colour) and hospitalisations (lighter colour) with the total number of each given in the top left hand corner of each panel. The black bars represent when the precautionary breaks take place in each instance. In this figure, the left column shows the results when GDP is fitted to the whole of 2020, whilst the right column shows the results when GDP is fitted from June to December 2020.

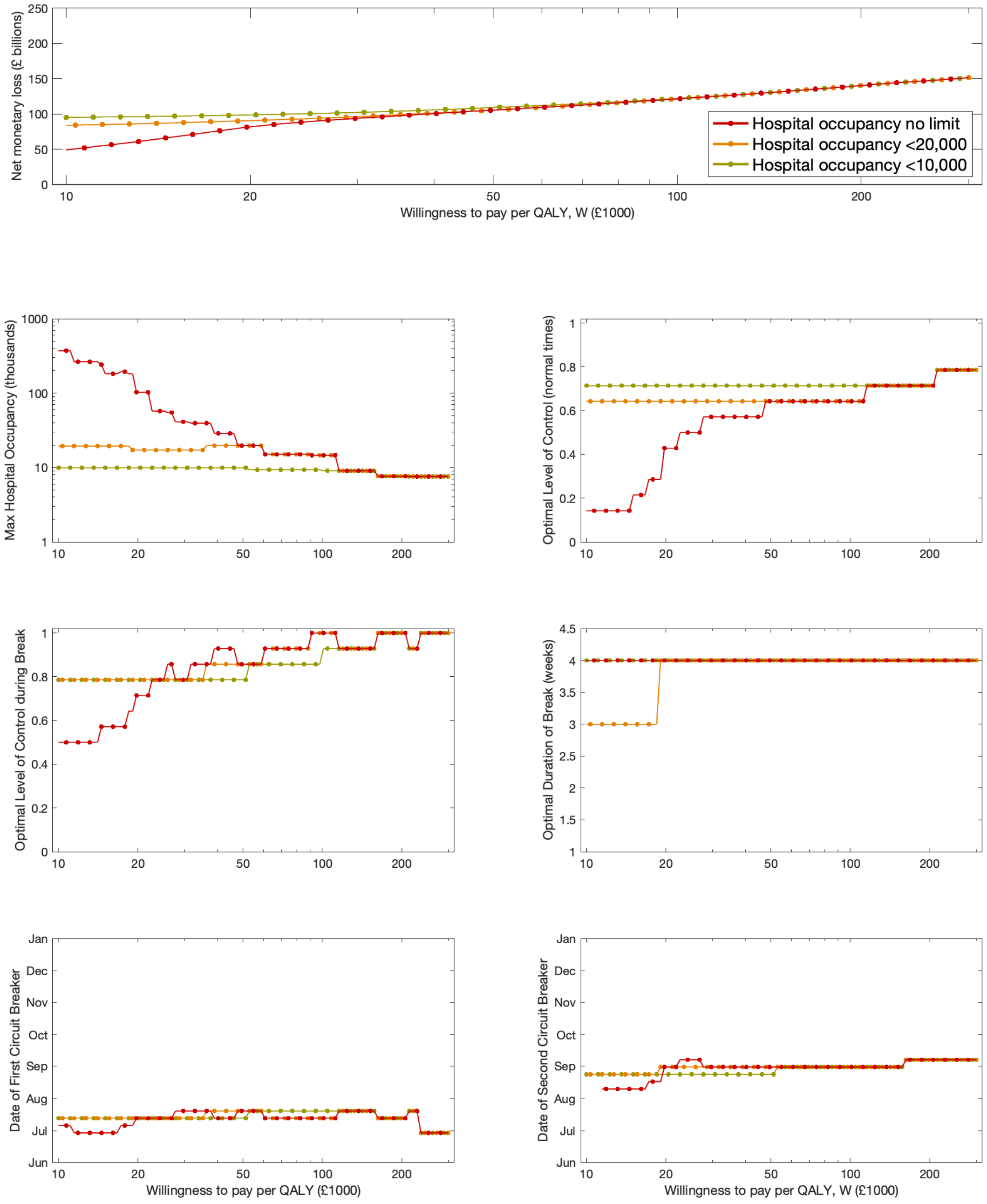


**Fig S5: Outcomes over a range of willingness to pay per QALY ($W$) values. (Top Row)** Net monetary loss (W $\times$ QALY loss + GDP loss) against different values of $W$ as the daily hospitalisation threshold varies (different colours). In rows two-four, we display the following measures for the optimal control strategy as the willingness to pay per QALY increases: **(second row, left panel)** maximum number of hospital admissions per day. **(second row, right panel)** the optimal level of intrinsic control outside lockdown; **(third row, left panel)** the optimal level of control within a precautionary break; **(third row, right panel)** the optimal duration of the lock-down in days; **(fourth row, left panel)** the optimal date of the first precautionary break; **(fourth row, right panel)** the optimal date of the second precautionary break. In this figure we fit to GDP for all 2020.

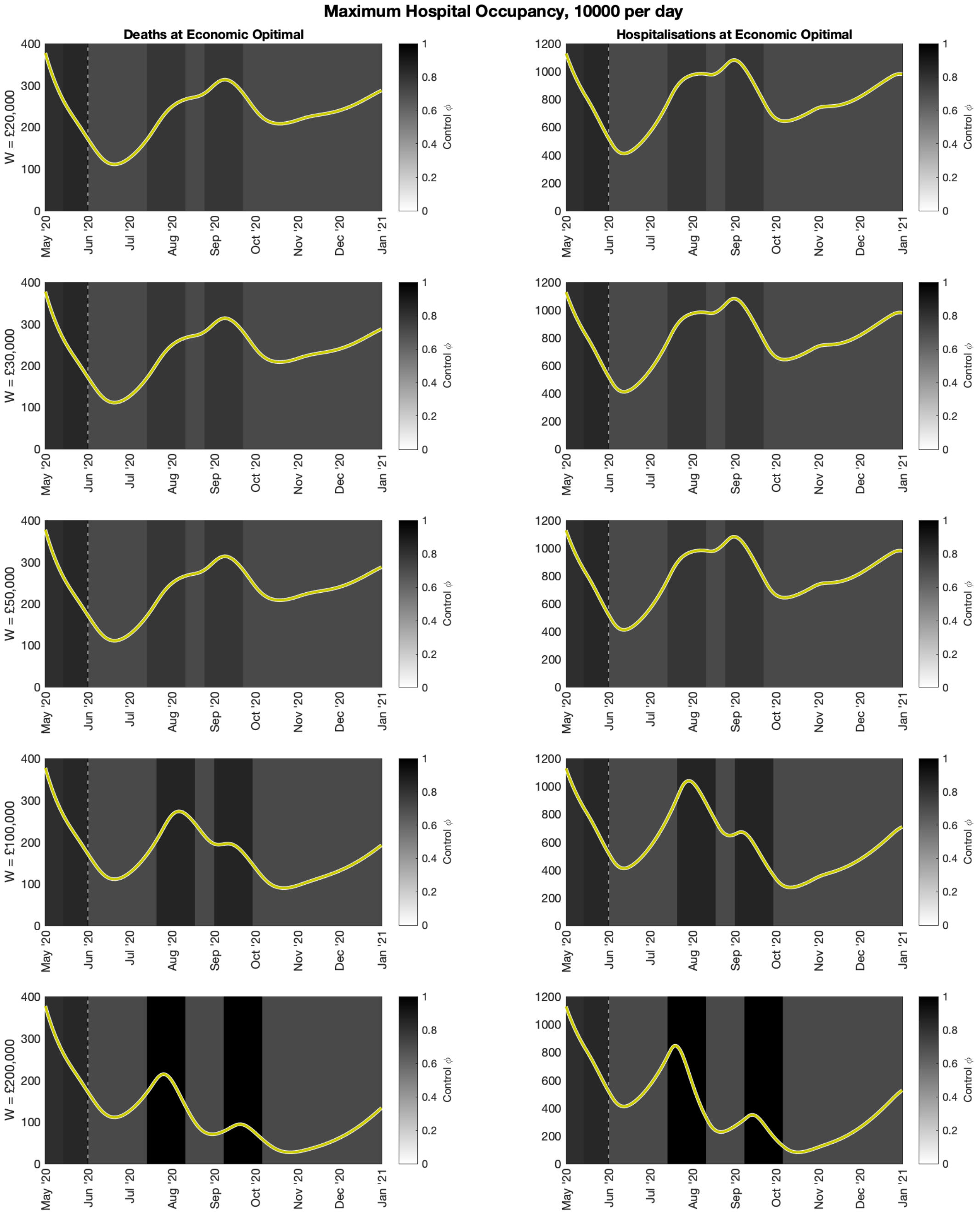


**Fig S6: Optimal control strategy with a maximum hospital occupancy threshold of 10000 individuals per day.** Daily deaths (left column) and daily hospital admissions (right column) for the optimal control strategy with a maximum hospital occupancy threshold of 10000 individuals per day, as the willingness to pay varies from £20,000 per QALY (top row) to £200,000 per QALY (bottom row). The shading in each figure shows the level of control occurring at each point in time, with simulations commencing from 1st June 2020 (vertical dashed line). The darker vertical bands from July 2020 onwards show the periods of time in which precautionary breaks should occur to minimise the overall loss, given the value of the willingness to pay. In this figure, we fit to GDP for all 2020.

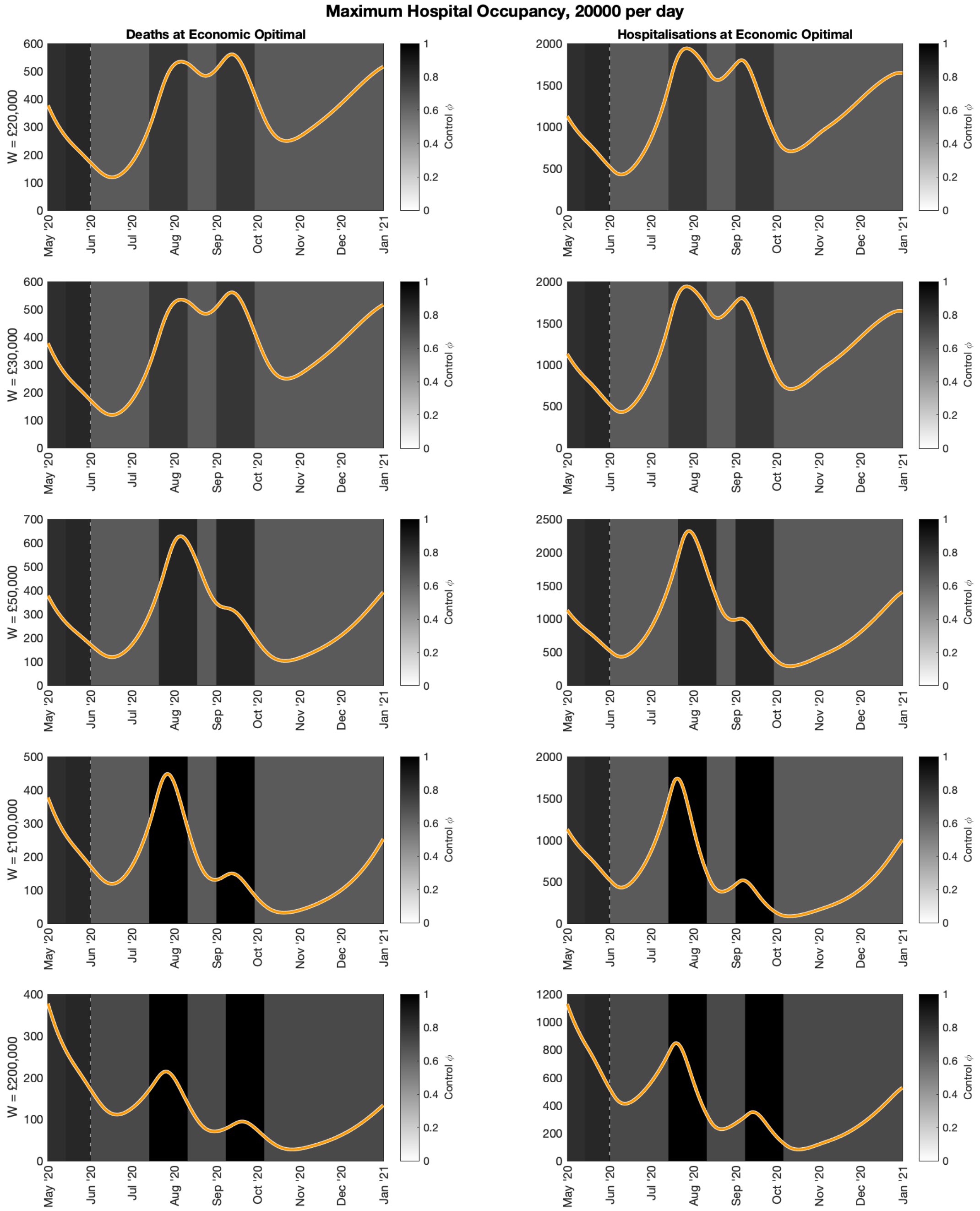


**Fig S7: Optimal control strategy with a maximum hospital occupancy threshold of 20000 individuals per day.** Daily deaths (left column) and daily hospital admissions (right column) for the optimal control strategy with a maximum hospital occupancy threshold of 20000 individuals per day, as the willingness to pay varies from £20,000 per QALY (top row) to £200,000 per QALY (bottom row). The shading in each figure shows the level of control occurring at each point in time, with simulations commencing from 1st June 2020 (vertical dashed line). The darker vertical bands from July 2020 onwards show the periods of time in which precautionary breaks should occur to minimise the overall loss, given the value of the willingness to pay. In this figure, we fit to GDP for all 2020.

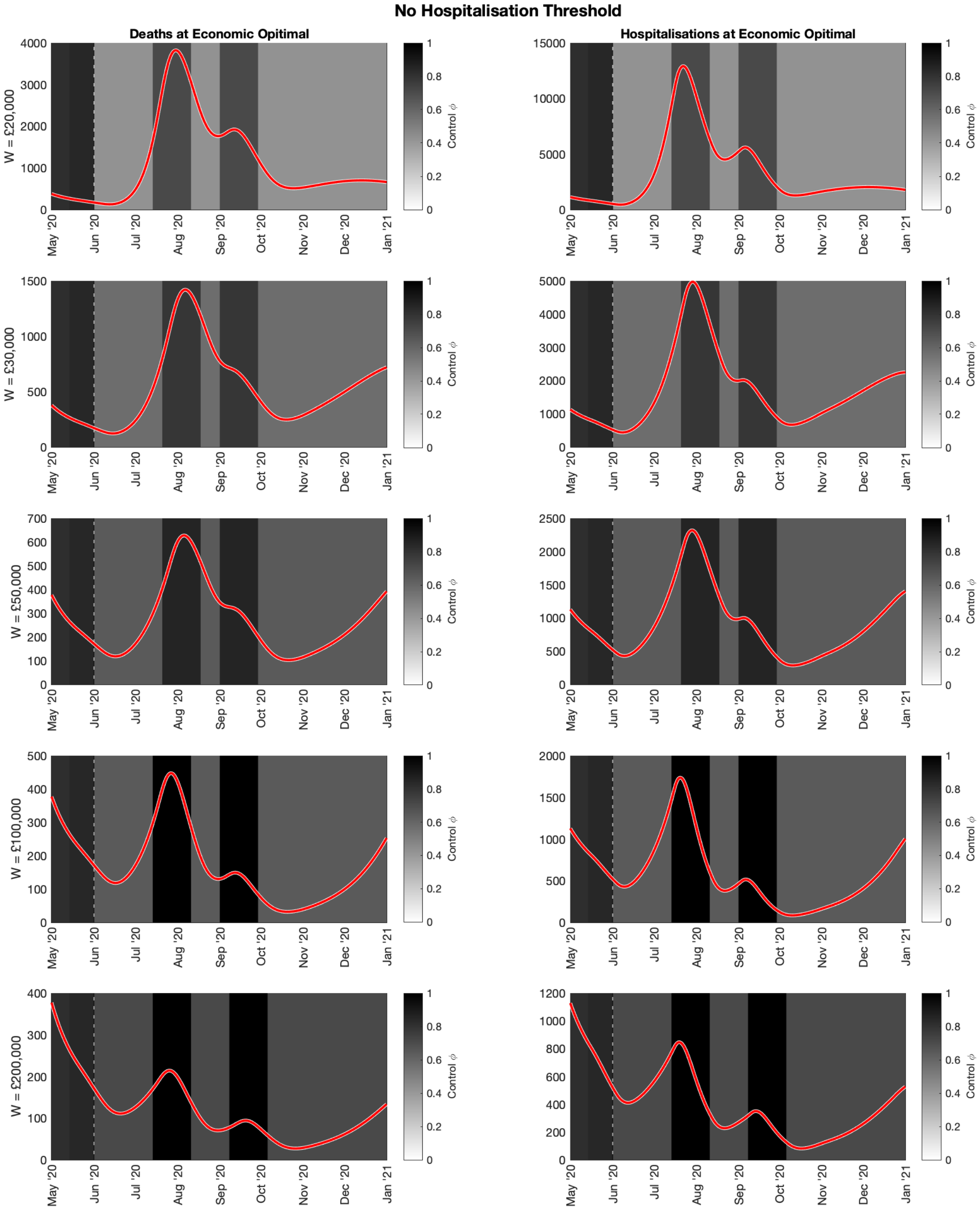


**Fig S8: Optimal control strategy with no maximum hospital occupancy threshold.** Daily deaths (left column) and daily hospital admissions (right column) for the optimal control strategy with no maximum hospital occupancy threshold, as the willingness to pay varies from £20,000 per QALY (top row) to £200,000 per QALY (bottom row). The shading in each figure shows the level of control occurring at each point in time, with simulations commencing from 1st June 2020 (vertical dashed line). The darker vertical bands from July 2020 onwards show the periods of time in which precautionary breaks should occur to minimise the overall loss, given the value of the willingness to pay. In this figure, we fit to GDP for all 2020.

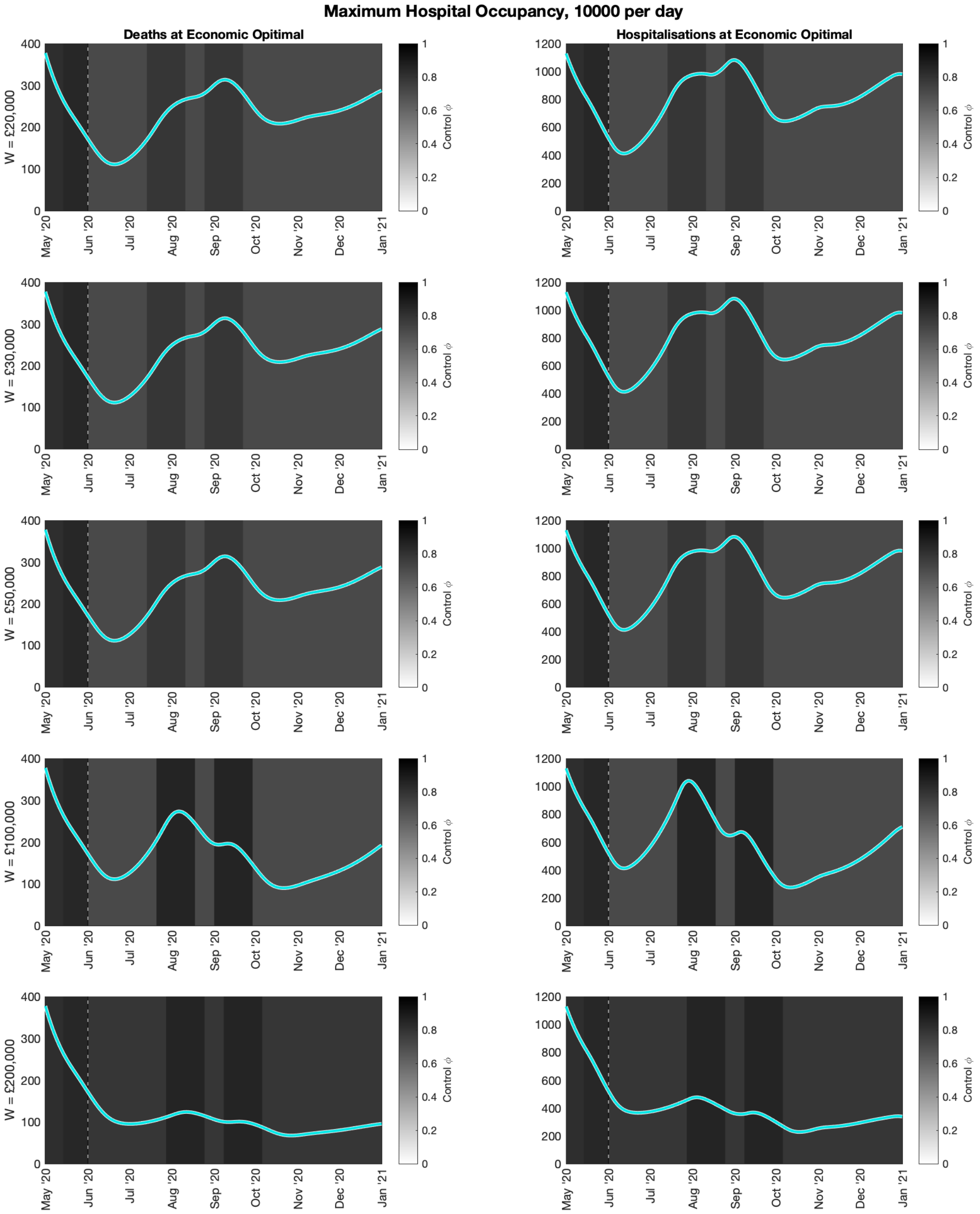


**Fig S9: Optimal control strategy with a maximum hospital occupancy threshold of 10000 individuals per day.** Daily deaths (left column) and daily hospital admissions (right column) for the optimal control strategy with a maximum hospital occupancy threshold of 10000 individuals per day, as the willingness to pay varies from £20,000 per QALY (top row) to £200,000 per QALY (bottom row). The shading in each figure shows the level of control occurring at each point in time, with simulations commencing from 1st June 2020 (vertical dashed line). The darker vertical bands from July 2020 onwards show the periods of time in which precautionary breaks should occur to minimise the overall loss, given the value of the willingness to pay. In this figure, we fit to GDP from June to December 2020.

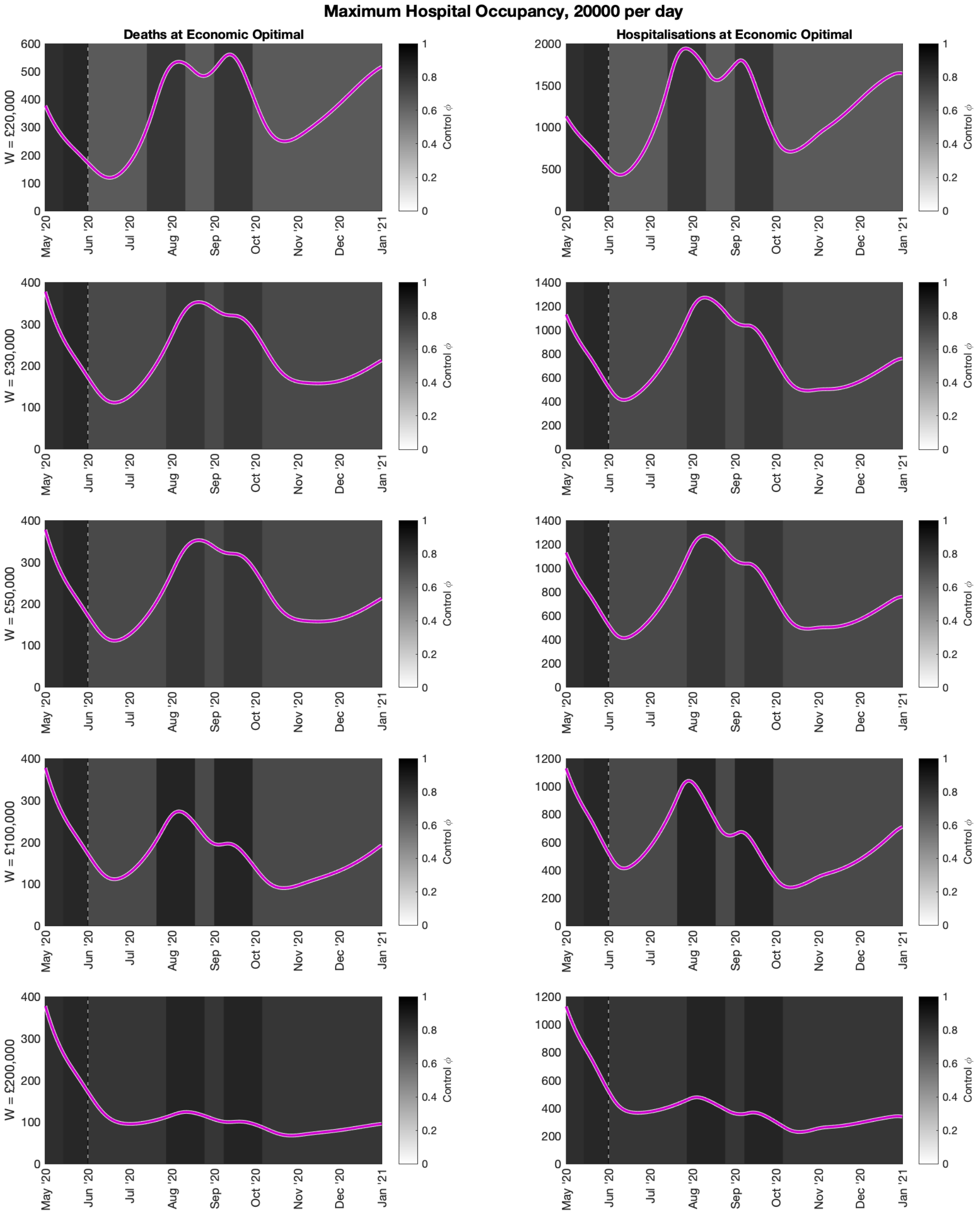


**Fig S10: Optimal control strategy with a maximum hospital occupancy threshold of 20000 individuals per day.** Daily deaths (left column) and daily hospital admissions (right column) for the optimal control strategy with a maximum hospital occupancy threshold of 20000 individuals per day, as the willingness to pay varies from £20,000 per QALY (top row) to £200,000 per QALY (bottom row). The shading in each figure shows the level of control occurring at each point in time, with simulations commencing from 1st June 2020 (vertical dashed line). The darker vertical bands from July 2020 onwards show the periods of time in which precautionary breaks should occur to minimise the overall loss, given the value of the willingness to pay. In this figure, we fit to GDP from June to December 2020.

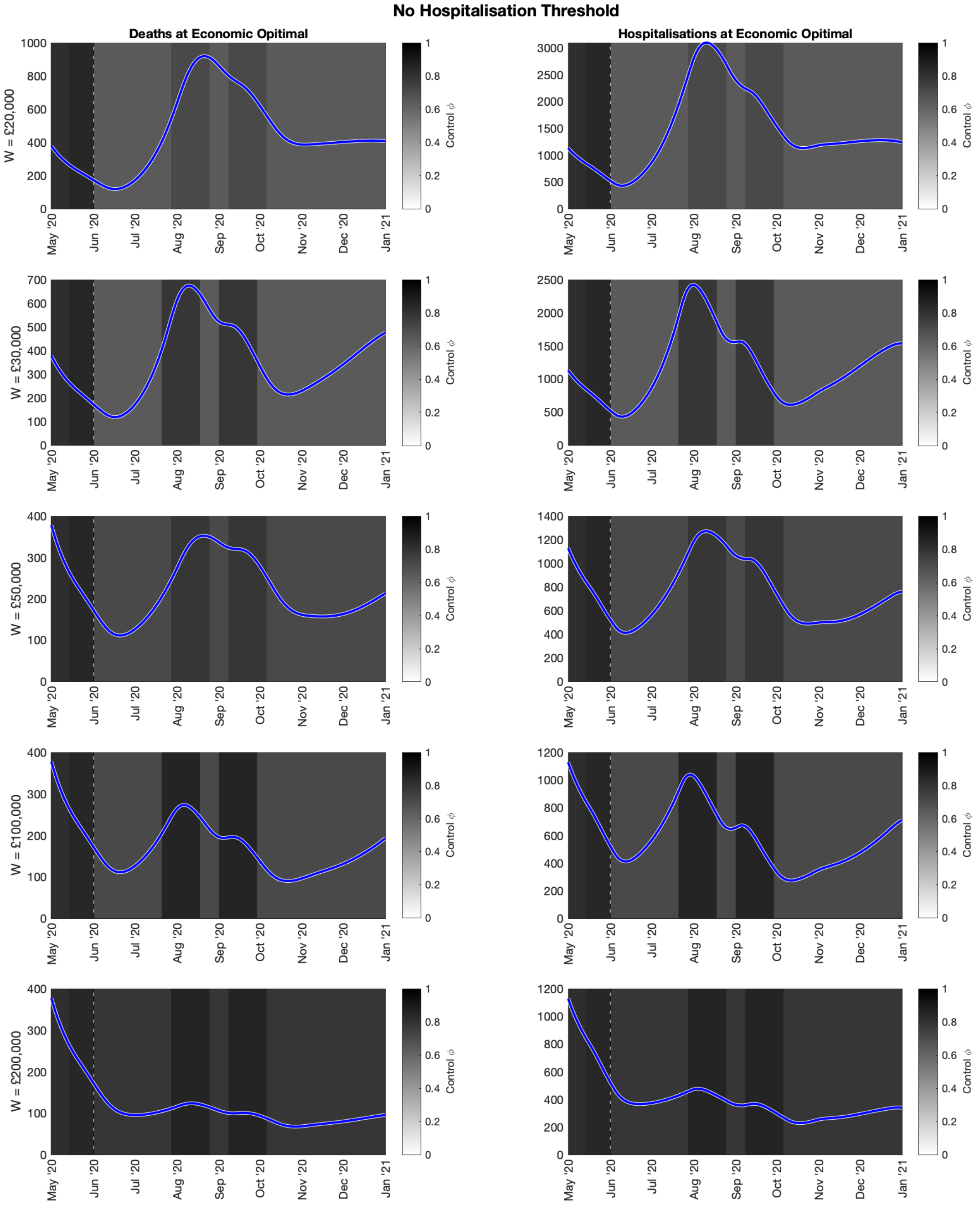


**Fig S11: Optimal control strategy with no maximum hospital occupancy threshold.** Daily deaths (left column) and daily hospital admissions (right column) for the optimal control strategy with no maximum hospital occupancy threshold, as the willingness to pay varies from £20,000 per QALY (top row) to £200,000 per QALY (bottom row). The shading in each figure shows the level of control occurring at each point in time, with simulations commencing from 1st June 2020 (vertical dashed line). The darker vertical bands from July 2020 onwards show the periods of time in which precautionary breaks should occur to minimise the overall loss, given the value of the willingness to pay. In this figure, we fit to GDP from June to December 2020.
